# An Approach for Open Multivariate Analysis of Integrated Clinical and Environmental Exposures Data

**DOI:** 10.1101/2021.06.30.21259727

**Authors:** Karamarie Fecho, Perry Haaland, Ashok Krishnamurthy, Bo Lan, Stephen A. Ramsey, Patrick L. Schmitt, Priya Sharma, Meghamala Sinha, Hao Xu

**Author notes:** Authors are listed alphabetically. Corresponding author: Karamarie Fecho, PhD, University of North Carolina at Chapel Hill, RENCI, 100 Europa Drive, Suite 540, Chapel Hill, North Carolina 27517.

## Abstract

The Integrated Clinical and Environmental Exposures Service (ICEES) provides regulatory-compliant open access to sensitive patient data that have been integrated with public exposures data. ICEES was designed initially to support dynamic cohort creation and bivariate contingency tests. The objective of the present study was to develop an open approach to support multivariate analyses using existing ICEES functionalities and abiding by all regulatory constraints. We first developed an open approach for generating a multivariate table that maintains contingencies between clinical and environmental variables using programmatic calls to the open ICEES application programming interface. We then applied the approach to data on a large cohort (N = 22,365) of patients with asthma or related conditions and generated an eight-feature table. Due to regulatory constraints, data loss was incurred with the incorporation of each successive feature variable, from a starting sample size of N = 22,365 to a final sample size of N = 4,556 (20.5%), but data loss was < 10% until the addition of the final two feature variables. We then applied a generalized linear model to the subsequent dataset and focused on the impact of seven select feature variables on asthma exacerbations, defined as annual emergency department or inpatient visits for respiratory issues. We identified five feature variables—sex, race, obesity, prednisone, and airborne particulate exposure—as significant predictors of asthma exacerbations. We discuss the advantages and disadvantages of ICEES open multivariate analysis and conclude that, despite limitations, ICEES can provide a valuable resource for open multivariate analysis and can serve as an exemplar for regulatory-compliant informatics solutions to open patient data, with capabilities to explore the impact of environmental exposures on health outcomes.

## INTRODUCTION

Interest in open access to and sharing of electronic health record (EHR) data has been growing in recent years, both among the medical research community and patient advocacy groups. The benefits of such an effort are perhaps best highlighted by the current coronavirus pandemic and the need to rapidly initiate research into the virus and its impacts on health, as well as share data and findings and develop a global response to this unprecedented health crisis. Large-scale Initiatives such as the Columbia Open Health Data (COHD) [1] and Medical Information Mart for Intensive Care (MIMIC) [2] are advancing efforts to reduce regulatory and institutional barriers surrounding access to EHR data, with the common goal of promoting research, while preserving patient privacy and maintaining institutional assurances. However, further efforts are required to truly leverage EHR data and apply the data to promote global human health and well-being.

As part of the Biomedical Data Translator (‘Translator’) program [3,4], funded by the National Center for Advancing Translational Sciences, we have developed a novel, regulatory-compliant, disease-agnostic framework and approach for openly exposing and exploring EHR data that have been integrated at the patient level with a variety of public exposures data: the Integrated Clinical and Environmental Exposures Service (ICEES). ICEES is accessible to anyone on the internet via an application programming interface (API). We have validated ICEES by replicating published research on asthma and related common pulmonary diseases [5–8]. We have extended ICEES to expose multi-institutional data on patients within UNC Health who are also participants within the Environmental Polymorphisms Registry at the National Institute of Environmental Health Sciences [7]. Moreover, as part of the Translator program, we have used ICEES to conduct a multi-institutional study, free of regulatory constraints, over the course of a five-day ‘hackathon’ [10]. We also have developed a tool for visualizing and exploring ICEES as a ‘knowledge graph’ of interconnected nodes [11].

While ICEES has demonstrated technical validity and scientific application, the service remains subject to federal and institutional constraints that, while necessary, limit the available functionalities. ICEES currently supports the ability to dynamically define cohorts and explore bivariate relationships between feature variables such as diagnoses, medications, and airborne pollutant exposures. Herein, we describe the development of a novel open approach that supports multivariate analysis using existing regulatory-compliant ICEES functionalities, while maintaining all federal and institutional regulations and preserving patient privacy. We apply the open multivariate approach to a driving use case on asthma, using a generalized linear model (GLM) to predict asthma exacerbations. Finally, we discuss the advantages and disadvantages of using ICEES for multivariate analysis.

## MATERIALS AND METHODS

All study procedures were approved by the Institutional Review Board at the University of North Carolina at Chapel Hill.

### Technical Overview

#### Open Multivariate Approach

ICEES is equipped with regulatory-compliant analytic capabilities that allow users to dynamically create cohorts and generate bivariate contingency tables with corresponding Chi Square statistics, probabilities, and frequencies. Motivated by our desire to develop more sophisticated multivariate analytic capabilities here we describe the development and application of an open approach to conduct multivariate analysis using the functionalities that are currently available in ICEES. In sum, the approach leverages the dynamic cohort creation capability in such a way as to maintain feature contingencies and generate a multivariate table, while remaining compliant with all federal and institutional regulations.

#### ICEES Integrated Feature Tables

Key to the design of ICEES is what we’ve termed ‘ICEES integrated feature tables’. The tables are designed as one-year ‘study periods’, in which each row represents an individual patient and each column header represents a feature variable. The tables contain integrated data on clinical data elements derived from patient EHRs and exposures data derived from a variety of public sources (e.g., United States Environmental Protection Agency airborne particulate exposures, US Department of Transportation roadway exposures, US Census Bureau American Community Survey socioeconomic exposures, North Carolina Department of Environmental Quality concentrated animal farming operations exposures and landfill exposures). The integration step is achieved using a complex custom software pipeline [8] and requires protected health information (PHI; i.e., geocodes and dates). As such, this step is conducted under a protocol that must be approved by an Institutional Review Board. After integration, however, all PHI elements are removed from the data according to §164.514(b) of the Health Insurance Portability and Accountability Act [12]. The data are then exposed via an open ICEES API [13] that adheres to the Translator Application Programming Interface (TRAPI) standards [14].

### Generation of Multivariate ICEES Integrated Feature Tables

We developed the general approach in the context of a driving application use case, in which we asked if there is a relationship between asthma exacerbations and the following demographic features, clinical characteristics, and environmental exposures: sex, race, prescriptions for prednisone, diagnosis of obesity, residential proximity to a major roadway or highway, residential density, and exposure to airborne particulates. These variables were selected on the basis of published studies, including our prior work [6,8, 9], which identified these variables as known or suspected to be related to asthma exacerbations. We focused on an existing ICEES cohort of UNC Health patients with asthma or related conditions (see [6] for details), and we considered the number of annual emergency department (ED) or inpatient visits for respiratory issues as the primary outcome measure and indicator of asthma exacerbations. We examined asthma exacerbations in year 2010, as this was the first year of data available for the cohort.

The seven features or independent variables and the primary outcome metric or dependent variable are defined and enumerated in **Table 1**.

**Table 1.** Feature variables used to generate multivariate table.

| Feature Variable | Variable Definition and Enumeration |
| --- | --- |
| TotalEDInpatientVisits | *Total number ED or inpatient visits for respiratory issue(s) over the 'study' period (0, 1, 2, 3, ...)* |
| Sex2 | *Male (0), Female (1)* |
| Race | *Caucasian, African American, Asian, Native Hawaiian/Pacific Islander, American/Alaskan Native, Other* |
| Prednisone | *One or more prescriptions for common medication for asthma-like conditions (1=Yes, 0=No)* |
| ObesityDx | *One or more diagnostic codes for obesity (1=Yes, 0=No)* |
| MaxDailyPM2.5Exposure_StudyMax | *US Environmental Protection Agency estimated maximum daily exposure to airborne particulate matter $\leq 2.5\text{-}\mu\text{m}$ in diameter ($\text{PM}_{2.5}$) over 'study' period, binned using pandas.cut (1, 2, 3, 4, 5)* |
| RoadwayDistanceExposure2 | *US Department of Transportation distance in meters from household to nearest roadway (1 = 0-49, 2 = 50-99, 3 = 100-149, 4 = 150-199, 5 = 200-249, 6 = $\geq 250$ meters)* |
| EstResidentialDensity | *US Census Bureau American Community Survey 2007–2011 estimated total population [block group], binned according to US Census Bureau definitions (1=rural [0,2500), 2=urban cluster [2500,50000), 3=urbanized area [50000,inf))* |

Abbreviations: $\text{PM}_{2.5}$ = particulate matter $\leq 2.5\text{-}\mu\text{m}$ in diameter.

While ICEES supports functionalities to examine the bivariate relationship between ED/inpatient visits for respiratory issues and each of the feature variables of interest, it does not directly support the application of multivariate statistical or machine learning models to examine relationships and interactions across multiple feature variables. To apply multivariate models, an eight-feature table was required, with each row representing an individual patient and each column header representing a distinct feature variable, with contingencies maintained across feature variables. To achieve this, we applied the ICEES dynamic cohort creation functionality and used nested bivariate contingencies to generate the requisite multivariate feature table. A visual overview of the approach is provided in **Figure 1**.

**Figure 1.**
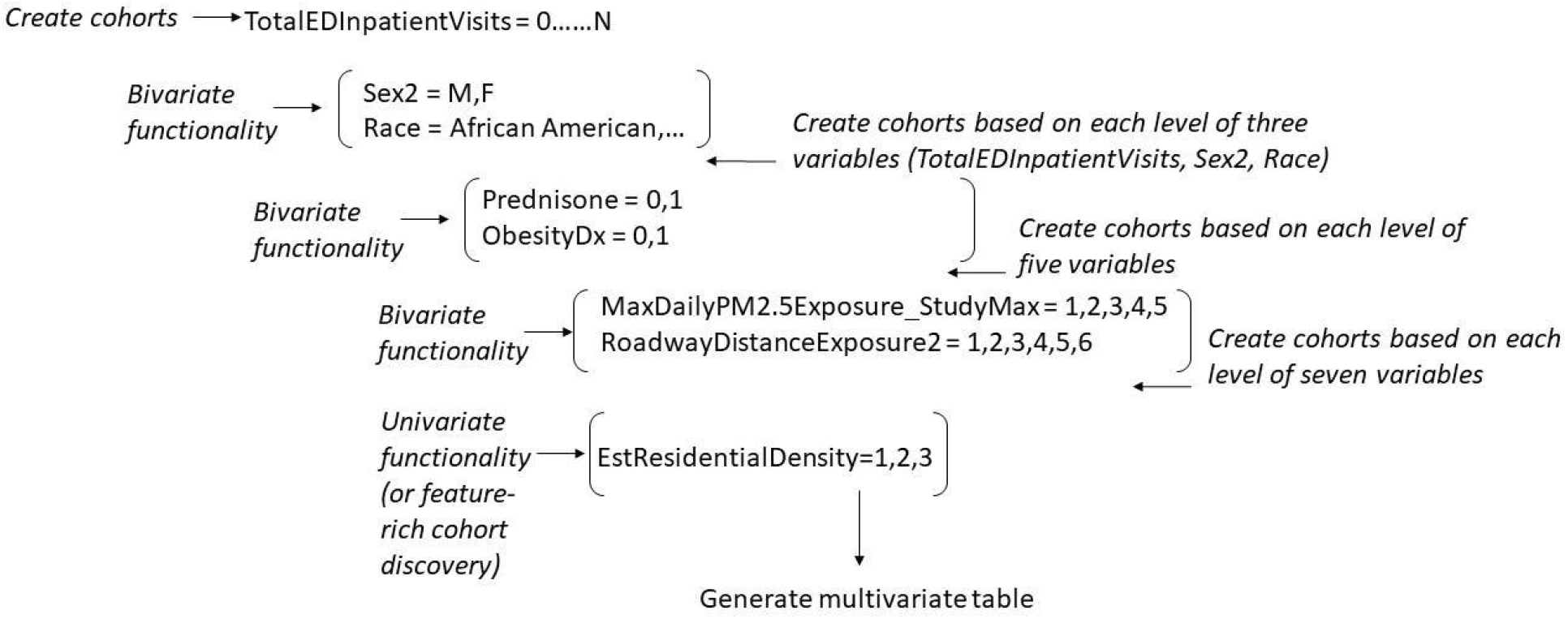
High-level overview of process for generating an ICEES multivariate table by application of dynamic cohort creation and nested bivariate contingencies. Levels or bins for each variable are defined in source documentation available from the OpenAPI and also accessible as an ICEES OpenAPI endpoint. See **Table 1** for the feature variable definitions and enumeration used in this study.

Specifically, we first selected the asthma cohort, table type (patient or visit), table version, and calendar year of interest as the input parameters. We then created separate cohorts for each level of the dependent variable (i.e., ED/inpatient visits for respiratory issues):

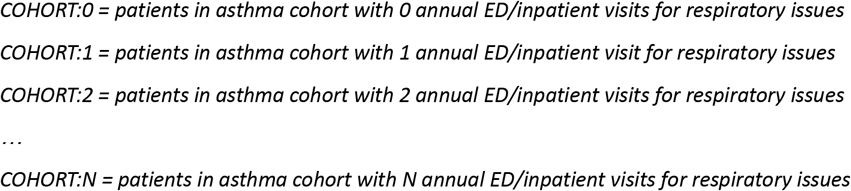

The boundary of COHORT:N is determined by both the underlying data (i.e., the maximum number of annual ED/inpatient visits reported for a patient in any given year) and the regulatory constraints imposed on the ICEES service, namely, that cohorts with <10 patients cannot be created, in which case, the service returns an error message indicating that the data do not exist or that the selected cohort consists of <10 patients. The practical implication of this regulatory constraint for the efforts described here was that a certain amount of data loss was incurred with each step in the process of generating a multivariate table. We quantified the data loss by comparing the size of the sum of each cohort or the number of rows for each intermediary table with the size of the overall sample.

The next step in the process for creating a multivariate table was to create a bivariate contingency table for each of the cohorts generated in the first step. In our example use case, we used *Sex2 x Race*. Because the contingencies between feature variables were maintained, we were then able to create a tri-variate table, with rows transformed to represent *N = 1 patient* (**Figure 2**).

**Figure 2.**
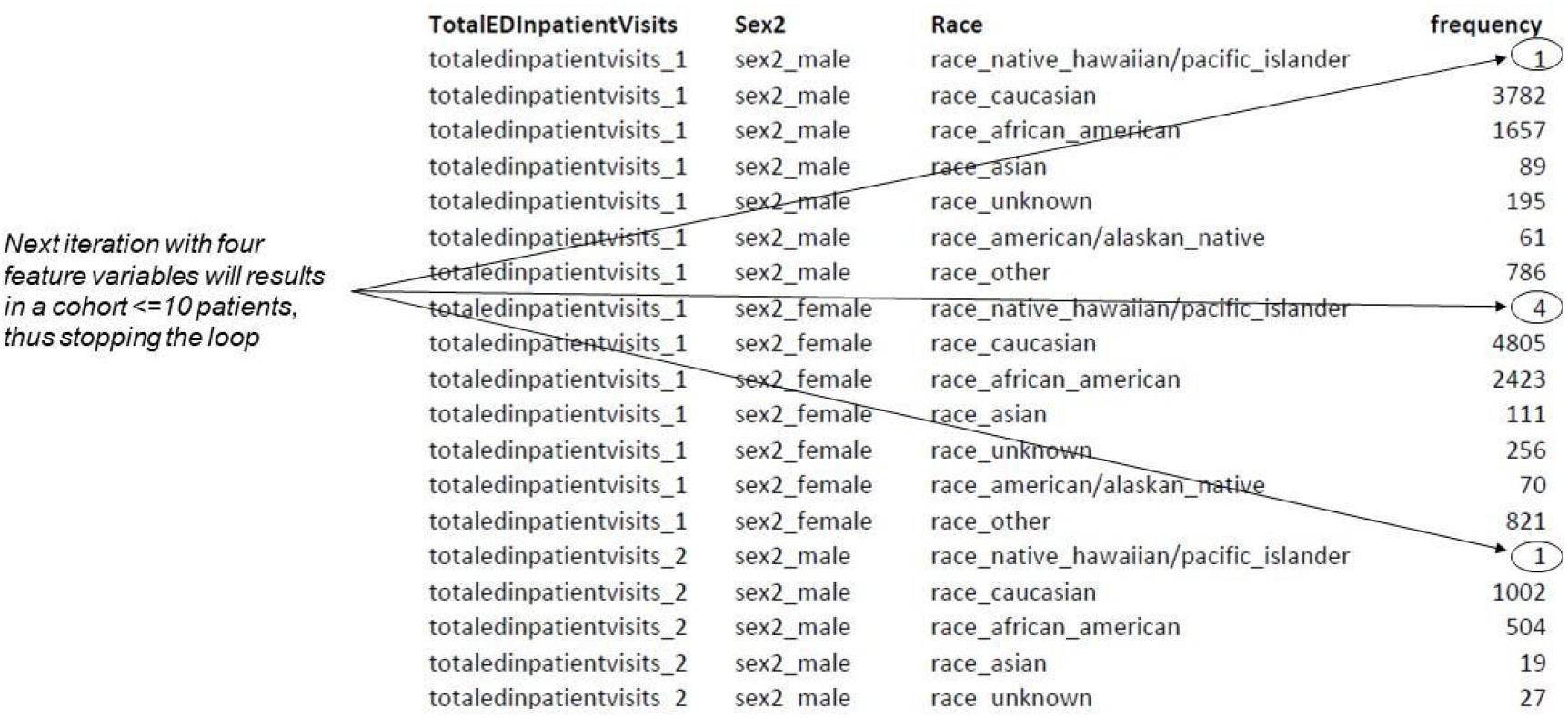
Example ICEES tri-variate table, with rows in aggregate form representing the number of patients sharing the characteristics defined in each column. Each row can thus be duplicated to represent *N = 1* patient.

The next step was to create cohorts for each combination of the three feature variables.

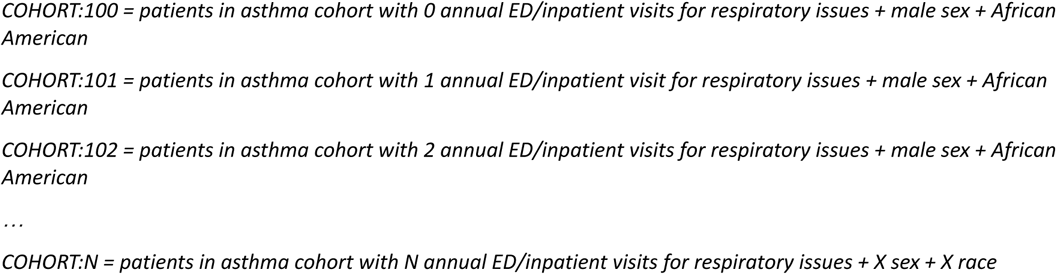

For each of the new cohorts, a second bivariate contingency table was generated. In our example, the association was for *Prednisone x ObesityDx*. The cohort creation and bivariate contingency table steps were then repeated for *MaxDailyPM2*.*5Exposure_StudyMax x RoadwayDistanceExposure2*. As we were interested in an odd number of independent variables, the final step that we applied to the data was to invoke the ICEES univariate functionality (also called feature-rich cohort discovery) to examine frequencies for *EstResidentialDensity*. Upon completion of this step, we then were able to generate an eight-feature multivariate table, with each row representing an individual patient (see Results). For interpretation purposes, and to minimize data loss, we categorized the dependent variable, *TotalEDInpatientVisits*, as 0, 1, … 9+.

### Application of Multivariate GLM

We developed a GLM algorithm using R to predict *TotalEDInpatientVisits* using the seven independent feature variables extracted in the ICEES multivariate table. Given that *TotalEDInpatientVisits* are counts and that the distribution was skewed to the right (i.e., few patients have frequent ED visits or hospital admissions for respiratory issues in any given year), we fit a negative binomial model to the data [15]. We also applied the Synthetic Minority Oversampling Technique (SMOTE) [16] to account for imbalances in the data because the frequencies for certain variables (e.g., *RoadwayDistanceExposure2*) were not evenly distributed across bins. The SMOTE approach augments the minority class in order to balance the data such that model performance accounts for cells with otherwise low frequencies. We examined both main effects and interactions, and then applied an analysis of variance (ANOVA) to the obtained GLM results, with α = 0.05.

## RESULTS

### Eight-feature Multivariate Table and Estimated Data Loss

We applied the ICEES open multivariate approach to generate an eight-feature multivariate table designed to support our driving application use case on the effects of select demographic variables, socio-economic exposures, and airborne pollutant exposures on asthma exacerbations (**Figure 3**).

**Figure 3.**
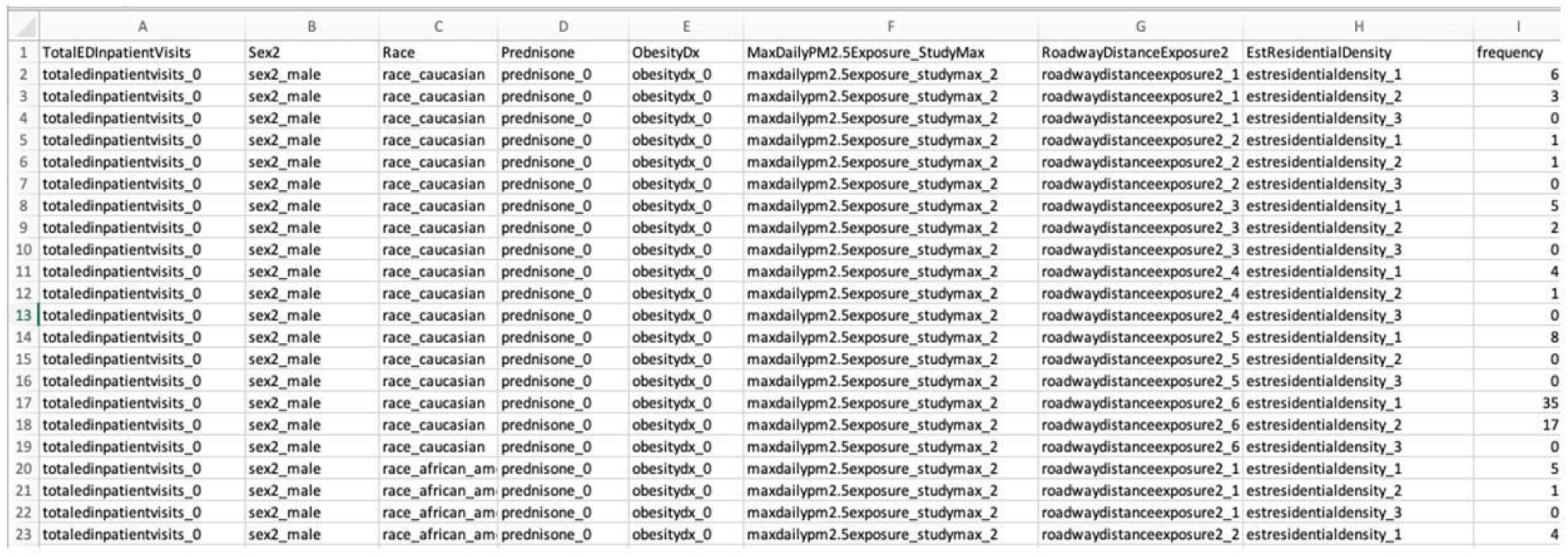
Excerpt from ICEES eight-feature multivariate table. The frequency column allows users to generate patient-level rows by, for instance, creating six separate rows for the features defined in row one and assigning a pseudo-identifier.

We then quantified the data loss that occurs with each step in the process by comparing the size of each recreated cohort with that of the initial cohort (**Table 2**). Data loss was incurred after the fourth independent variable was incorporated and increased to 57.1% with the incorporation of the seventh and final independent variable.

**Table 2.** Quantification of data loss with ICEES open multivariate approach.^*^

| Feature Variable Added† | Total ICEES Rows (N) | Maximum Possible Rows (N) | Missing Rows (N) | Missing Rows/Maximum Possible Rows (%) |
| --- | --- | --- | --- | --- |
| Starting sample size | 22365 | N/A | N/A | N/A |
| Sex2 | 22365 | 22365 | 0 | 0 |
| Race | 22365 | 22365 | 0 | 0 |
| Prednisone | 22365 | 22365 | 0 | 0 |
| ObesityDx | 22208 | 22361 | 153 | 0.68 |
| MaxDailyPM2.5Exposure_StudyMax | 15861 | 17390 | 1529 | 8.79 |
| RoadwayDistanceExposure2 | 5022 | 8262 | 3240 | 39.2 |
| EstResidentialDensity | 4556 | 10615 | 6059 | 57.1 |

\*Starting sample size before filtering for patients who were active in the study period (calendar year 2010): N = 163302
†Feature variables were added in the order listed, following the schema shown in **Figure 1**.

### Application Use Case Results

We applied a GLM algorithm to the resultant multivariate table and asked the following specific use-case question: *are sex, race, prescriptions for prednisone, diagnosis of obesity, residential proximity to a major roadway or highway, residential density, and/or exposure to airborne particulates predictive, either independently or by interaction, of asthma exacerbations?*

We found significant main effects of *Race, Prednisone, ObesityDx, MaxDailyPM2*.*5Exposure_StudyMax*, and *Sex2* (**Table 3**). Several two- and three-way interactions also were significant. Among two-way interactions, *Prednisone* showed significant interactions with *Sex2, ObesityDx, MaxDailyPM2*.*5Exposure_StudyMax*, and *RoadwayDistanceExposure2*. A significant *Sex2* x *ObesityDx* effect also was apparent. Among three-way interactions (data not shown), *ObesityDx* x *Sex2* x *Race, ObesityDx* x *Sex2* x *Prednisone*, and *ObesityDx* x *Race* x *Prednisone* were significant. Higher-level interactions were not significant.

**Table 3.**
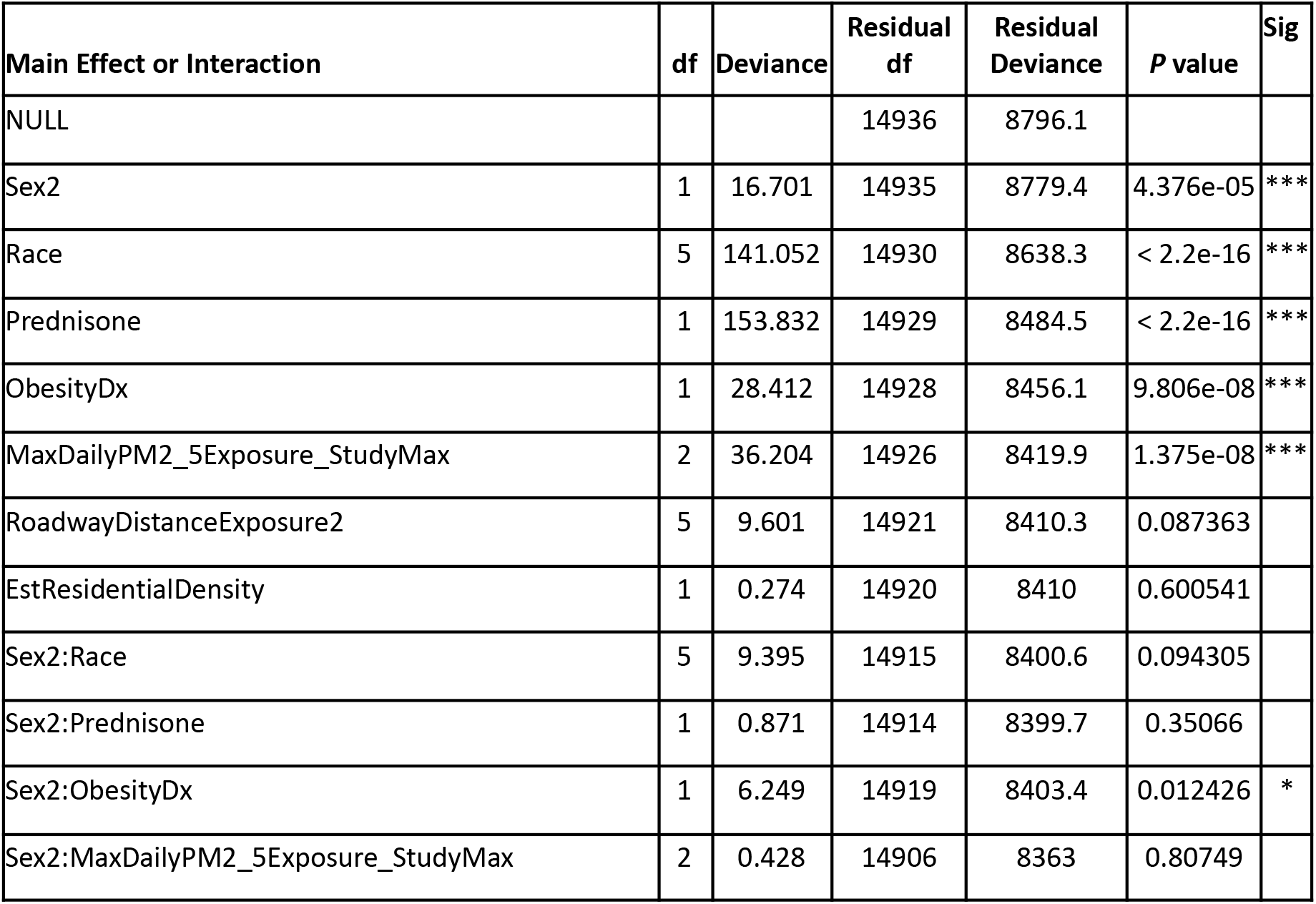
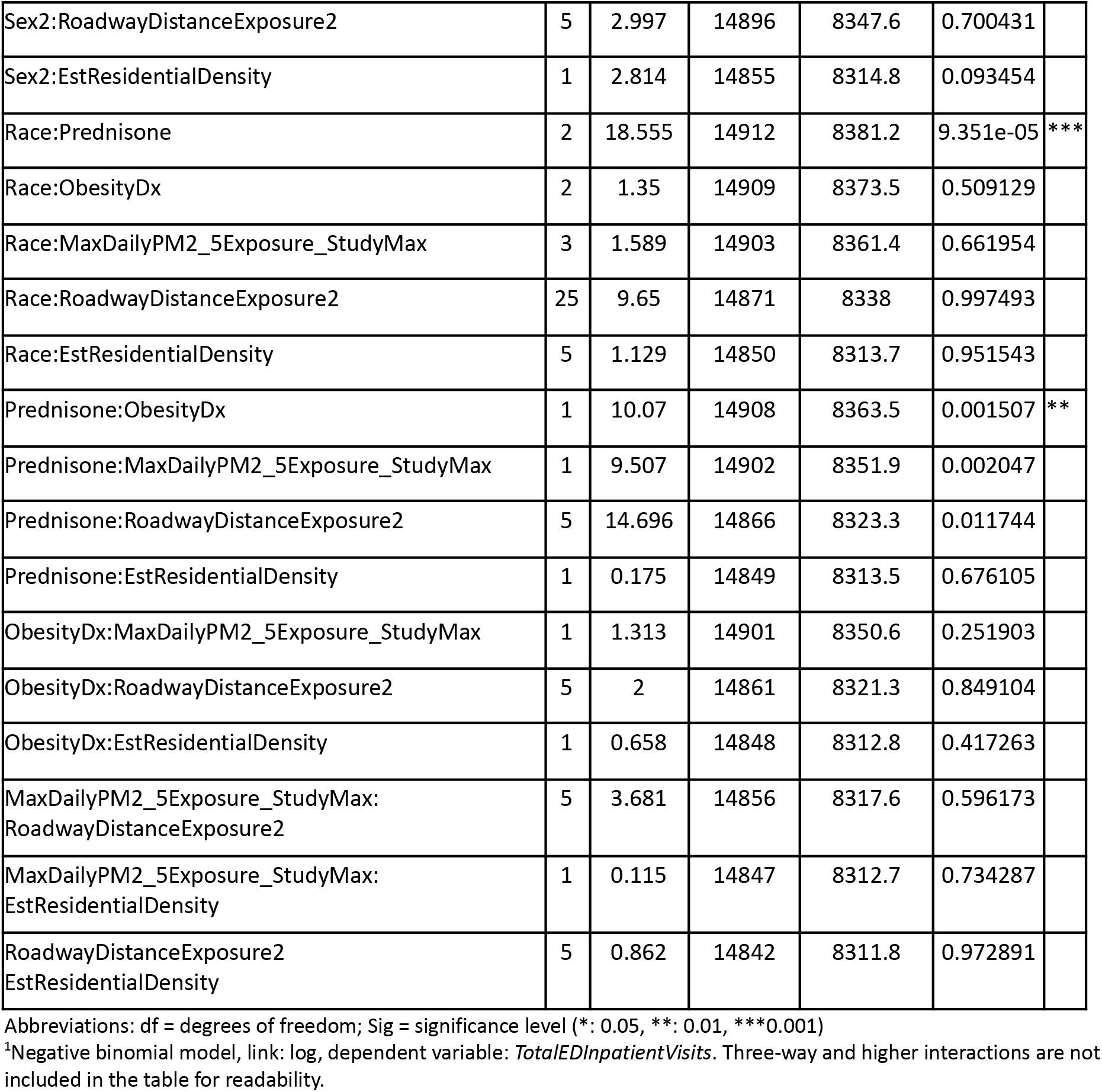
ANOVA results for GLM model with main effects and 2-way interactions.^1^

| Main Effect or Interaction | df | Deviance | Residual df | Residual Deviance | P value | Sig |
| --- | --- | --- | --- | --- | --- | --- |
| NULL |  |  | 14936 | 8796.1 |  |  |
| Sex2 | 1 | 16.701 | 14935 | 8779.4 | 4.376e-05 | *** |
| Race | 5 | 141.052 | 14930 | 8638.3 | < 2.2e-16 | *** |
| Prednisone | 1 | 153.832 | 14929 | 8484.5 | < 2.2e-16 | *** |
| ObesityDx | 1 | 28.412 | 14928 | 8456.1 | 9.806e-08 | *** |
| MaxDailyPM2_5Exposure_StudyMax | 2 | 36.204 | 14926 | 8419.9 | 1.375e-08 | *** |
| RoadwayDistanceExposure2 | 5 | 9.601 | 14921 | 8410.3 | 0.087363 |  |
| EstResidentialDensity | 1 | 0.274 | 14920 | 8410 | 0.600541 |  |
| Sex2:Race | 5 | 9.395 | 14915 | 8400.6 | 0.094305 |  |
| Sex2:Prednisone | 1 | 0.871 | 14914 | 8399.7 | 0.35066 |  |
| Sex2:ObesityDx | 1 | 6.249 | 14919 | 8403.4 | 0.012426 | * |
| Sex2:MaxDailyPM2_5Exposure_StudyMax | 2 | 0.428 | 14906 | 8363 | 0.80749 |  |
| Sex2:RoadwayDistanceExposure2 | 5 | 2.997 | 14896 | 8347.6 | 0.700431 |  |
| Sex2:EstResidentialDensity | 1 | 2.814 | 14855 | 8314.8 | 0.093454 |  |
| Race:Prednisone | 2 | 18.555 | 14912 | 8381.2 | 9.351e-05 | *** |
| Race:ObesityDx | 2 | 1.35 | 14909 | 8373.5 | 0.509129 |  |
| Race:MaxDailyPM2_5Exposure_StudyMax | 3 | 1.589 | 14903 | 8361.4 | 0.661954 |  |
| Race:RoadwayDistanceExposure2 | 25 | 9.65 | 14871 | 8338 | 0.997493 |  |
| Race:EstResidentialDensity | 5 | 1.129 | 14850 | 8313.7 | 0.951543 |  |
| Prednisone:ObesityDx | 1 | 10.07 | 14908 | 8363.5 | 0.001507 | ** |
| Prednisone:MaxDailyPM2_5Exposure_StudyMax | 1 | 9.507 | 14902 | 8351.9 | 0.002047 |  |
| Prednisone:RoadwayDistanceExposure2 | 5 | 14.696 | 14866 | 8323.3 | 0.011744 |  |
| Prednisone:EstResidentialDensity | 1 | 0.175 | 14849 | 8313.5 | 0.676105 |  |
| ObesityDx:MaxDailyPM2_5Exposure_StudyMax | 1 | 1.313 | 14901 | 8350.6 | 0.251903 |  |
| ObesityDx:RoadwayDistanceExposure2 | 5 | 2 | 14861 | 8321.3 | 0.849104 |  |
| ObesityDx:EstResidentialDensity | 1 | 0.658 | 14848 | 8312.8 | 0.417263 |  |
| MaxDailyPM2_5Exposure_StudyMax:<br>RoadwayDistanceExposure2 | 5 | 3.681 | 14856 | 8317.6 | 0.596173 |  |
| MaxDailyPM2_5Exposure_StudyMax:<br>EstResidentialDensity | 1 | 0.115 | 14847 | 8312.7 | 0.734287 |  |
| RoadwayDistanceExposure2<br>EstResidentialDensity | 5 | 0.862 | 14842 | 8311.8 | 0.972891 |  |

Abbreviations: df = degrees of freedom; Sig = significance level (\*: 0.05, \*\*: 0.01, \*\*\*0.001)
<sup>1</sup>Negative binomial model, link: log, dependent variable: *TotalEDInpatientVisits*. Three-way and higher interactions are not included in the table for readability.

## DISCUSSION

We demonstrated the ability to programmatically use existing regulatory-compliant ICEES functionalities (i.e., dynamic cohort creation and bivariate contingencies) to generate a multivariate integrated feature table. Importantly, we developed and applied a GLM model to the resultant multivariate table and identified five feature variables—*Prednisone, Race, ObesityDx, Sex*2, *MaxDailyPM2*.*5Exposure_StudyMax*—as significant predictors of *TotalEDInpatientVisits*.

Importantly, our application findings are in agreement with the published literature. For instance, prednisone is commonly prescribed for the treatment of acute asthma exacerbations in patients who are non-responsive to first-line treatments such as inhaled albuterol [17]. Female sex, obesity, and African American race have previously been identified as variables that contribute to asthma exacerbations. For example, Greenblatt et al. [18] found that female sex and obesity (among other variables) significantly increased the odds of asthma exacerbations. Our prior work [8] and that of others [19] have found a significant association between African American race and increased risk of asthma exacerbations. Finally, exposure to airborne particulate matter is a well-established risk factor for asthma and asthma exacerbations. For example, a study by Requia et al. [20] found a significant association between a two-year increase of 10 µg/m^3^ PM_2.5_ in 117 regions in Canada and increased risk in the incidence of asthma. Mirabelli et al. [21] likewise found a significant association between exposure to PM_2.5_ and risk of asthma. Exposure to major roadways or highways is often used as a proxy for airborne particulate exposure. Indeed, several groups have demonstrated an increase in asthma exacerbations among patients residing in close proximity to a major roadway or highway [22,23]. We did not identify major roadway/highway exposure as a significant predictor of asthma exacerbations. As our patient population is largely rural (unpublished observation), we speculate that exposure to major roadways or highways may not be of primary relevance to asthma exacerbations.

While we have validated the ICEES open multivariate approach described here, several considerations are worthy of discussion. First, ICEES multivariate tables must be created in the context of a driving use case question, with a dependent variable identified and defined as the starting point for the overall approach. While this is not a limitation *per se*, it is a consideration that users should take into account.

Second, while the ICEES multivariate analytic approach is openly available, the ICEES service itself is subject to regulatory constraints that limit the amount of data that can be accessed and the types of analyses that can be performed. Specifically, cohorts ≤ 10 patients cannot be created. The impact of this constraint is that a certain amount of data loss will be incurred whenever a cohort is created that has less than 10 patients. We are developing a theoretical framework to estimate data loss with different combinations of variables. For instance, suppose the most favorable case, namely, that each feature has only two values and that patients are divided equally among the possible values. Let there be *k* features in the query. Then, the ultimate cohort, say *C(k-1)*, must have at least 10 subjects to be included in the query output. The penultimate cohort, say *C(k-2)*, must have >=4*10 patients as a minimum. Therefore, the root cohort, say *C(1)*, must have >= (4^(k-2))*10 patients as a minimum. If there are eight features, then |*C(1)*| >= (4^6)*10 = 4096*10 = 40,960 patients are required at minimum to ensure that there is no data loss under the simplest, most favorable assumptions above. We plan to develop a technical approach for presenting this information to users so that they can apply the multivariate approach in an informed manner.

Third and related to the above consideration, the choice and order of variables influence the open multivariate approach and the final sample size. For instance, a variable that has many missing values or multiple levels will by definition decrease the final sample size. In some cases, the impact of this limitation can be quite large. For example, ICEES currently exposes data on genomic variants, but the data are available for only a minor subset of patients, and so incorporating genomic features from ICEES into a multivariate analysis is not realistic.

Fourth, the data loss that is inherent in the ICEES open multivariate approach may impact model quality. Consider that the final table has one row for every combination of the selected features. The frequency returned for any row in the table for which any previous cohort in the query was less than 10 will be returned as zero, regardless of the true value, but we know that the true value cannot be greater than If low-frequency rows are randomly distributed across the selected features, then we could assume that the query process may reduce the precision of the model results, but it would not introduce bias. In contrast, if low-frequency rows are not randomly distributed across the selected features, then we may introduce bias into our models, which will systematically affect the accuracy of model results and may lead to spurious conclusions. We are exploring approaches to anticipate and minimize bias.

Regardless of the limitations, we believe that the ICEES open multivariate approach provides a unique, regulatory-compliant service, with broad application. We are now comparing GLM model robustness and results with the API output versus the underlying data. We are also developing additional multivariate models such as random forest and causal inference. Finally, we are expanding the service to support additional use cases, including primarily ciliary dyskinesia and other rare respiratory disorders, drug-induced liver injury, coronavirus infection, and rare disease phenotypes.

## Data Availability

The ICEES API openly exposed clinical data that have been integrated with environmental exposures data

## Acknowledgments

The authors thank Dr. David Peden and Dr. Shepherd Schurman for providing expertise on the use case. The authors also thank their colleagues within the Biomedical Data Translator Consortium for their intellectual input and support for the work described herein.

## Competing Interests

The authors declare no competing interests.

## Funding Support

This project was funded with awards from the National Center for Advancing Translational Sciences, National Institutes of Health [OT3TR002020, OT2TR003430, UL1TR002489, UL1TR002489-03S4] and the Clinical Research Branch, Intramural Research Program of the National Institute of Environmental Health Sciences, National Institutes of Health [ZID ES103354-01].

## Notes

### Competing Interest Statement

The authors have declared no competing interest.

### Author Declarations

All study procedures have been approved by the Institutional Review Board at the University of North Carolina at Chapel Hill (protocol #16-2978)

